# Type 1 Dolichocolon as a Potential Anatomic Co-Morbidity in Pediatric Perianal Crohn’s Disease

**DOI:** 10.64898/2025.12.10.25341790

**Authors:** Richard Kellermayer, Réka G. Szigeti, Marla Sammer, Adam M. Vogel, Harland Winter

## Abstract

**Background:** Perianal Crohn’s disease (PCD) represents one of the most severe and refractory forms of pediatric inflammatory bowel disease (IBD). Constipation and colonic redundancy, particularly type 1 dolichocolon (T1-DC), may increase distal rectosigmoid pressure, and exacerbate perianal pathology. We hypothesized that T1-DC is more common in children with PCD than in those with uncomplicated ileocolonic Crohn’s disease (CD) or non-IBD controls.

**Methods:** We retrospectively analyzed 20 consecutive pediatric PCD cases (penetrating [B3p] or inflammatory [B1p]) and compared them with 20 patients with non-complicated ileocolonic CD (L3/B1) and 30 non-IBD trauma controls. DC type was determined radiographically using established criteria, focusing on T1- and T2-DC. Constipation history was abstracted from medical records under IRB-approved protocols.

**Results:** DC was significantly more prevalent in PCD than in ileocolonic CD or controls (p < 0.001), primarily due to T1-DC. The associations persisted (p<0.03) in PCD patients without a history of constipation.

**Conclusions:** Rectosigmoid redundancy (T1-DC) may represent an underrecognized anatomic co-morbidity in pediatric PCD, contributing to increased distal pressure and susceptibility to perianal complications. Identification of T1-DC could inform surgical decision-making and postoperative management. Targeted approaches—such as segmental resection during stoma reversal, structured bowel regimens, physical activity, and pelvic-floor biofeedback—may help reduce recurrence risk. Prospective studies are needed to define the mechanistic role of colonic redundancy in the pathogenesis of PCD.

## Introduction

Perianal Crohn’s or Crohn disease (PCD) in children is one of the most severe and disabling forms of inflammatory bowel disease (IBD). Characterized by fistulas, abscesses (penetrating disease), strictures as well as chronic ulceration, fissures, and inflamed tags (inflammatory disease) of the perineum, perianal disease is often refractory to medical treatment and may require a surgical procedure, such as a diverting ostomy in up to 40% of patients [1].

Constipation is often an overlooked comorbidity in pediatric Crohn’s disease (CD) that may occur prior to the diagnosis [2], or in some patients be a presenting symptom along with abdominal distension [3].

In patients with PCD, anatomic factors, such as colonic redundancy or dolichocolon [4], may be associated with constipation [5,6], extend transit time, [7] and increase fecal retention. Our recent work suggested that dolichocolon (DC) could influence pediatric ulcerative colitis phenotype [8]. These new observations suggest that constipation and colonic redundancy might complicate the management of PCD. In this study, we compared the prevalence of DC in PCD versus Crohn’s disease limited to the ileum and colon.

## Methods

### IRB/IACUC Approval

The study was conducted under institutional IRB-approved protocols (H-43969, H-50062) of Baylor College of Medicine.

### Description of Participants

We identified 20 consecutive pediatric cases and classified them according to Paris classification of CD [9] with perianal penetrating (B3p: abscess or fistula identified either physically or radiographically at diagnosis or during the disease course) or inflammatory (B1p: large, inflamed tags or fissures) PCD in a discovery cohort large tertiary care children’s hospital.

Inclusion criteria included no prior major abdominal surgery (except for appendectomy or cholecystectomy) and an age at diagnosis greater than 7 years (i.e. cases with very early onset IBD [VEO-IBD] were excluded). A history of constipation prior to or during the disease was recorded. The PCD patients were compared to 20 patients with uncomplicated L3/B1 ileocolonic Crohn’s disease without a change in the phenotype during a follow-up period for at least a year, and no history of treated constipation. Thirty (30) non-IBD controls (identified through electronic medical record search) were also included, who underwent abdominal CT scans for blunt abdominal trauma and had no documented constipation or recurrent abdominal pain in their electronic medical records.

DC has been traditionally divided into three types: cranial displacement of the sigmoid loop relative to the iliac crests (Type 1: T1-DC), caudal displacement of the transverse colon below the iliac crests (Type 2: T2-DC), and redundant loops at the hepatic or splenic flexures (Type 3)[5] Due to ambiguity in defining Type 3 DC, we have historically limited diagnostic consideration in pediatric cases to T1 and T2 DC, and selectively recorded Type 3. We used this approach in the current study to counteract bias since the iliac crest line represents a clear landmark for T1- and T2-DC.

DC prevalence was analyzed as a dichotomous variable using Fisher’s exact test (GraphPad Prism, Dotmatics, Boston, MA, USA). Group sizes were determined arbitrarily given the exploratory nature of this discovery study. Statistical significance was defined as p < 0.05.

## Results

Table 1 summarizes our findings regarding DC in PCD. DC was significantly (p<0.001) more common in PCD than in non-complicated, inflammatory ileocolonic (L3) CD (p=0.0008) or in trauma controls (p=0.0002) (Supplementary Figure 1). T1-DC accounted for this significant variation (Table 1 and Figure 1A and B). The increased prevalence of T1-DC was also observed in PCD patients without a recorded history of constipation in the EMR (“non-constipated” PCD; NC-PCD, compared to L3 (p=0.0239) and control (p=0.0271) patients as well (Figure 1C).

**Table 1.** DC prevalence with subtype distribution between the patient groups examined. CD: Crohn’s disease, CTRL (n=30): otherwise healthy, blunt force abdominal trauma patients (these patients had CT with contrast performed as trans-sectional abdominal imaging as opposed to the other groups who had MRI enterography or CT performed as standard of care), L3 CD: patients (n=20) with uncomplicated ileocolonic CD. PCD: perianal Crohn’s disease (n=20), N: number of patients, NC: no history of constipation. stdev: standard deviation. Age is in years of age when the trans-sectional imaging was performed. Bold: p ≤ 0.05 compared to L3 CD and Ctrl. P-values were calculated using Fisher’s exact test (GraphPad Prism, Dotmatics, Boston, MA, USA).

| Patient groups | PCD | L3 CD (NC) | CTRL (NC) |
| --- | --- | --- | --- |
| N | 20 [NC: 7] | 20 | 30 |
| Age (mean); +/-stdev | 13.4; +/- 3 | 12.7; +/- 2.2 | 12.4; +/- 3 |
| DC* (% of N) | **15 (75) [4 (57)]** | 3 (15) | 5 (17) |
| Type 1 only | **12 (60) [4 (57)]** | 2 (10) | 4 (13) |
| Type 2 only | 2 (10) | 1 (5) | 1 (3) |
| Type 1 and 2 | 1 (5) | 0 | 0 |
| Type 1 alone and with<br>Type 2 | 13 (65) | 2 (10) | 4 (13) |
| Type 2 alone and with<br>Type 1 | 3 (15) | 1 (5) | 1 (4) |
| Type 3 only (% of N) | 4 (20) | 6 (30) | 9 (30) |

**Figure 1.**
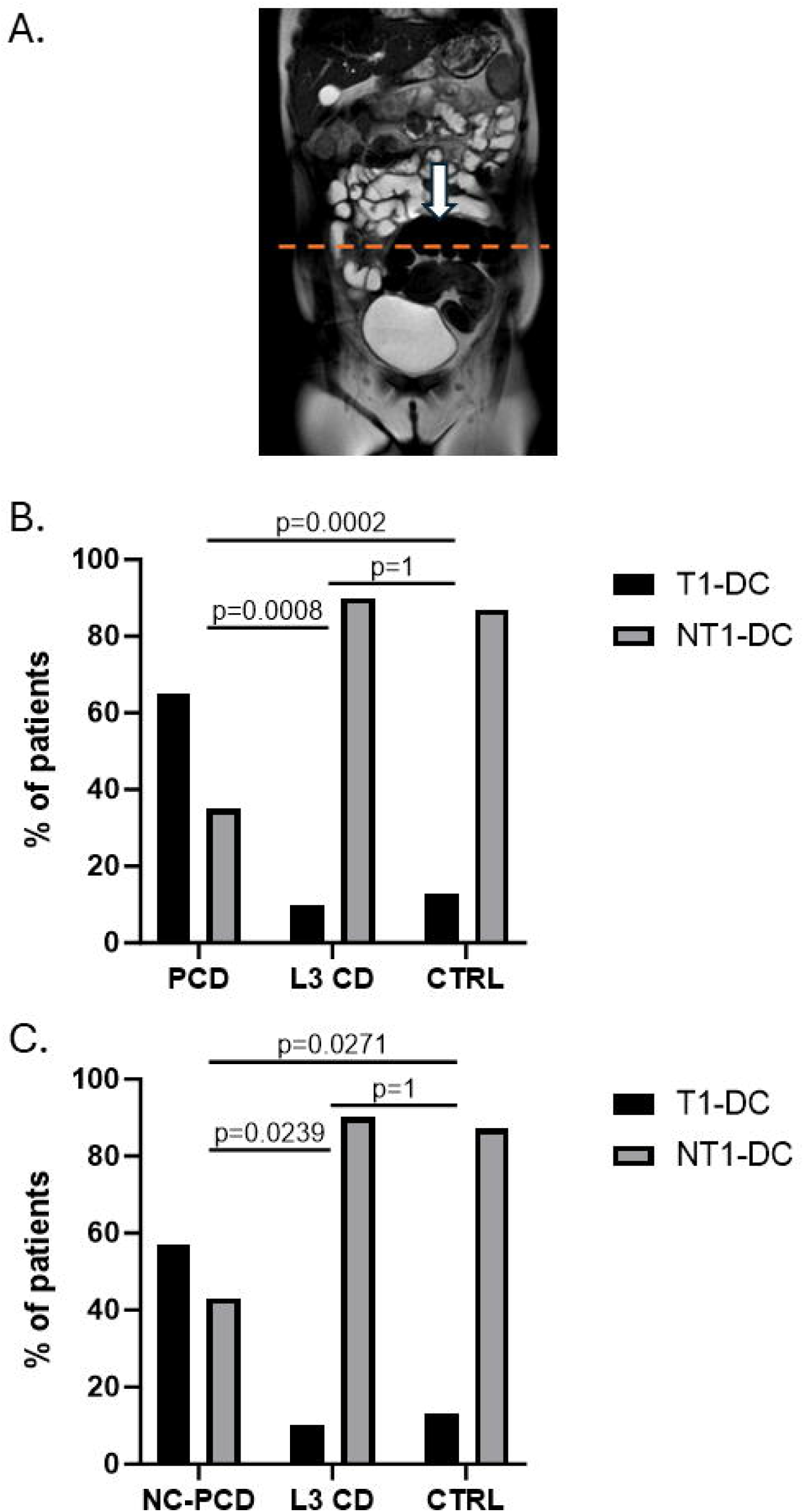
Type 1 Dolichocolon (T1-DC) in Pediatric Patients with Perianal Crohn’s disease (PCD, n=20). **A**. Representative case of T1-DC in an a young (more than 7 years of age) female patient with PCD at presentation. Note the redundant sigmoid colon (white arrow) emerging above the iliac crest line (dashed orange line). **B**. T1-DC was significantly more common in the PCD group compared to both control groups. **C**. This was true for those PCD patients who had no constipation history (NC-PCD) as well. P values were calculated in GraphPad Prism (Dotmatics, 225 Franklin Street, Floor 26 Boston, MA 02110, USA) by Fischer’s exact testing. CD: Crohn’s disease, CTRL (n=30): otherwise healthy, blunt force abdominal trauma patients, L3 CD: patients (n=20) with uncomplicated ileocolonic CD. NT1-DC: no Type 1 DC present.

## Discussion

This study is the first to highlight the potential complicating role of T1-DC in PCD. It is important to emphasize that penetrating (fistulizing) complications of Crohn’s disease (CD) almost always occur at or proximal to areas of stricturing disease [10], underscoring the role of increased luminal pressure in fistula formation. We speculate that the anal sphincter and the ileocecal valve effectively recreate the stricture-driven mechanism of increased luminal pressure, thereby helping to explain why the perianal region and terminal ileum are the most common sites for penetrating complications. Notably, increased anal resting pressure and rectal sensitivity have been observed in CD patients even in the absence of proctitis or perianal disease.[11]

Along with elevated anal resting pressure, our findings suggest that rectosigmoid redundancy (T1-DC) may play a significant role in the development of PCD, especially in pediatric patients. We believe that congenital DC, combined with genetic [12], microbial [13], and environmental [14] susceptibility factors, may be a co-morbidity in the development of PCD (Figure 2). Withholding stools in patients with sigmoid redundancy may predispose to the development of PCD. Repeated withholding and rectosigmoid fecal retention may further sustain or promote T1-DC (Figure 2).

**Figure 2:**
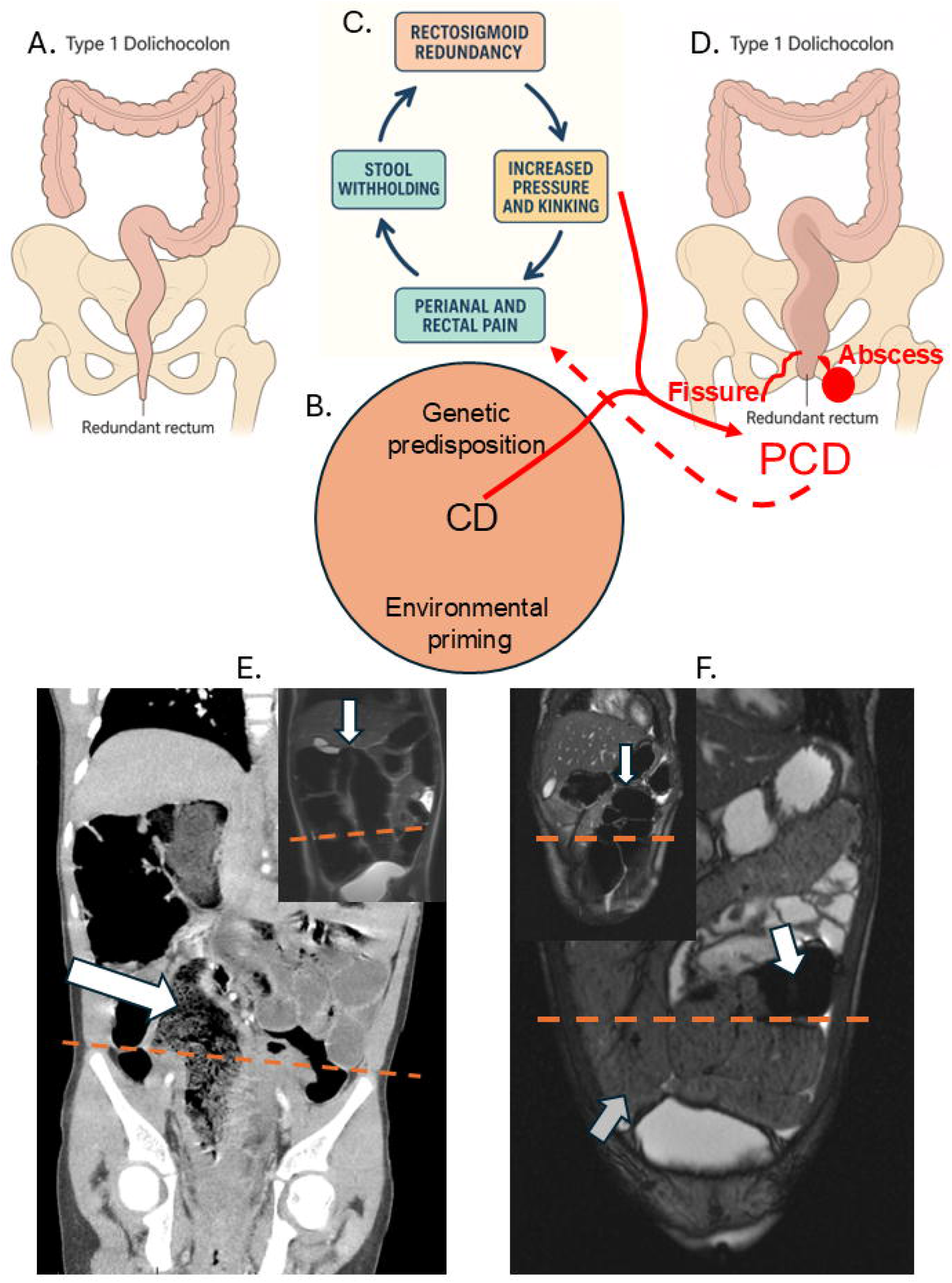
Dolichocolon in pediatric perianal Crohn’s disease. Schematic representation of the proposed pathomechanism of perianal Crohn’s disease (PCD), highlighting the proposed involvement of congenital Type 1 dolichocolon (A) in conjunction with genetic and environmental susceptibility factors (B) in pathogenesis. The vicious cycle of stool withholding (C) is aggravated by the progression of perianal penetrating and inflammatory disease in the context of Crohn’s disease (D). This process exacerbates pain-related withholding and rectosigmoid fecal retention, which further contributes to rectosigmoid elongation and redundancy. Panels E and F show MRI enterography images demonstrating Type 1 dolichocolon (DC) in two patients with very early-onset (age < 6 years) severe PCD prior to fecal diversion (low colostomy). White arrows indicate Type 1 DC with stool retention in the larger images, and the most proximal edge of the rectosigmoid junction in the insets. Orange dashed lines mark the iliac crest (IC) levels. The grey arrow in panel F highlights the transverse colon extending significantly below the IC line, consistent with Type 2 dolichocolon and meeting criteria for combined Type 1 and Type 2 DC in this case.

In this study, we specifically excluded patients with very early-onset inflammatory bowel disease (VEO-IBD) and perianal Crohn’s disease (PCD) because the North American Society for Pediatric Gastroenterology, Hepatology, and Nutrition considers VEO-IBD a distinct clinical entity that requires specialized diagnostic and management strategies, including genetic and immunologic assessments, due to its unique pathophysiology and treatment implications. [15] Nonetheless, VEO-IBD cases can offer valuable insights into the underlying mechanisms of IBD, particularly regarding genetic factors that may also influence later-onset disease. To support our hypothesis of T1-DC involvement in the disease process, we randomly selected two patients with VEO (age < 6 years at diagnosis), severe PCD, who underwent diverting colostomy. Profound T1-DC was found on their pre-diversion MRI enterography images (Figure 2E and F). These cases highlight the probable early developmental origins of T1-DC and suggest that, within the genetic and environmental context of CD, DC may contribute to a perianal phenotype in a patient-specific manner.

As with all retrospective studies, this work has inherent limitations. To minimize selection bias, we included all consecutive cases meeting the inclusion criteria across groups. Although blinding of observers for DC classification was not feasible, potential assessment bias was mitigated by applying strict anatomical criteria for T1- and T2-DC and excluding the less well-defined T3-DC from analysis. Despite relatively small group sizes, the observed association between T1-DC and PCD was statistically robust and clinically meaningful.

PCD occurs in approximately 14–33% of pediatric patients and 12–26% of adults presenting with Crohn’s disease [1,16-18]. In pediatric cohorts, the cumulative incidence rises to 28–45% over 10–30 years of follow-up [1,19]. In adults, the 10-year cumulative incidence is about 19%, with a lifetime risk up to 26% [17,20]. The proportion of pediatric patients with PCD who ultimately require surgical intervention, including a diverting ostomy, is approximately 40% [1]. The risk of surgery is higher in those with severe or complex perianal disease, rectal involvement, or refractory symptoms despite biologic therapy [1,16,21].

Most children who undergo fecal diversion for refractory PCD either remain diverted or ultimately require a permanent stoma due to relapses or persistent disease activity following attempts to restore bowel continuity. In a meta-analysis of more than 700 PCD cases managed with fecal diversion, only 29% achieved successful stoma reversal, and 31% of those subsequently required re-diversion [22]. It remains unclear whether patients with T1-DC or colonic redundancy who undergo diverting ostomy might benefit from resection of the elongated colonic segment at the time of stoma reversal, potentially reducing the risk of perianal disease recurrence. Adjunctive measures such as intensive bowel regimens, regular physical activity [23], and biofeedback therapy [24] to manage dolichocolon—by minimizing distal colonic pressure and fecal stasis—may further enhance postoperative outcomes and lower the likelihood of disease relapse.

## Data Availability

All data produced in the present study are available upon reasonable request to the authors

## Acknowledgements

R.K. was supported by the Gutsy Kids Fund, graciously maintained by Brock Wagner, the Klaasmeyers, the Frugonis, and other generous donor families. HSW received research support from The Hasso Serrano Foundation, Martin Schlaff, the Diane and Dorothy Brooks Foundation.

## Notes

Conflict of interest: None related to this manuscript

Conflict of interest: HSW: Consultant: Abbvie, Janssen Pharmaceuticals, Pfizer, Nestle, Women’s Wellness Foundation, Autism Research Institute, Pediatric IBD Foundation.

### Competing Interest Statement

HSW: Consultant: Abbvie, Janssen Pharmaceuticals, Pfizer, Nestle, Women's Wellness Foundation, Autism Research Institute, Pediatric IBD Foundation.

### Funding Statement

R.K. is a member of the Texas Medical Center, Digestive Disease Center funded by the NIH (NIDDK P30 DK 56338 Center for Gastrointestinal Development, Infection and Injury).

