## Supplemental Figure 1 for "Type 1 Dolichocolon as a Potential Anatomic Co-Morbidity in Pediatric Perianal Crohn’s Disease"

### Slide 1
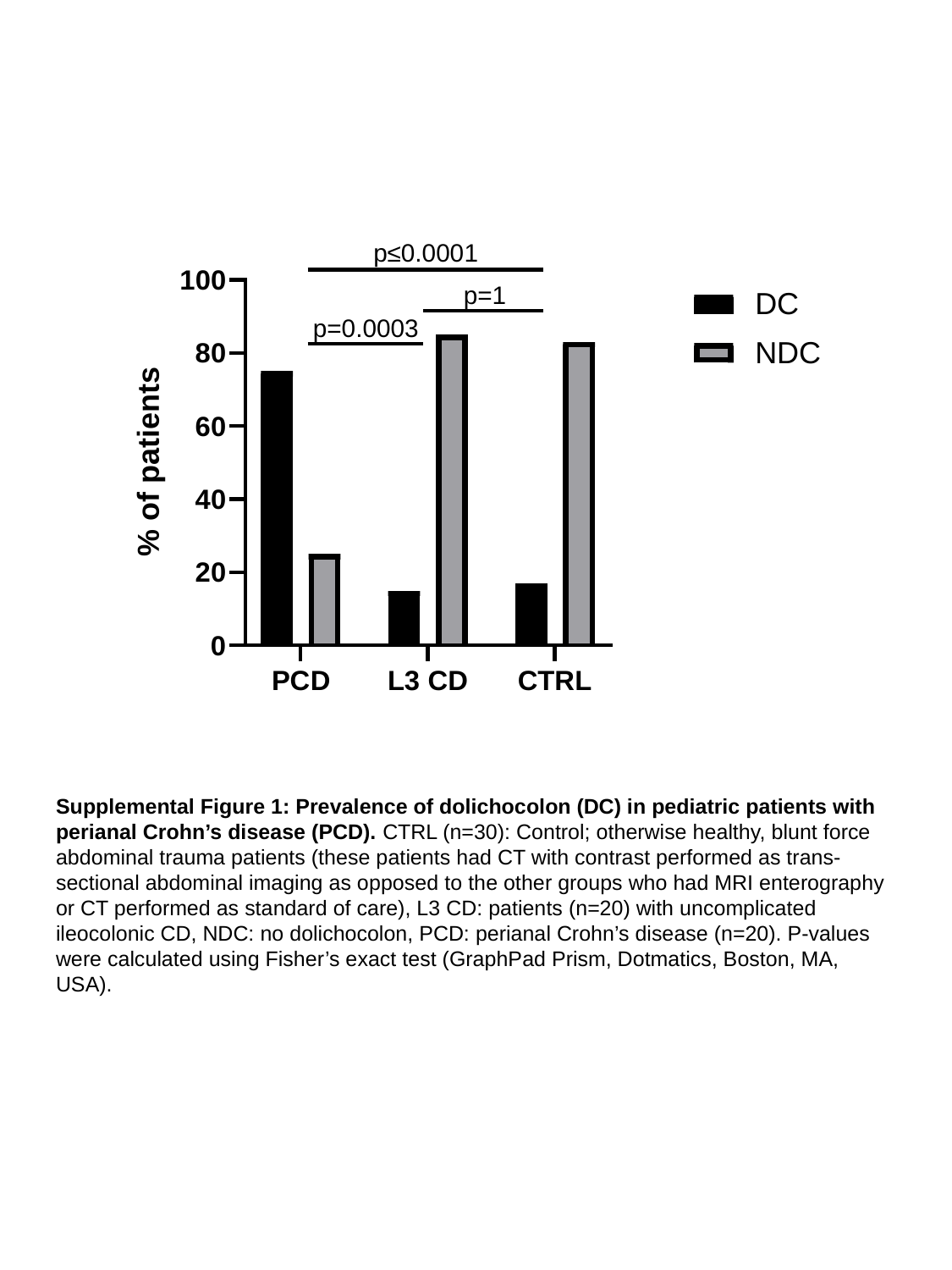

Supplemental Figure 1: Prevalence of dolichocolon (DC) in pediatric patients with perianal Crohn’s disease (PCD). CTRL (n=30): Control; otherwise healthy, blunt force abdominal trauma patients (these patients had CT with contrast performed as trans-sectional abdominal imaging as opposed to the other groups who had MRI enterography or CT performed as standard of care), L3 CD: patients (n=20) with uncomplicated ileocolonic CD, NDC: no dolichocolon, PCD: perianal Crohn’s disease (n=20). P-values were calculated using Fisher’s exact test (GraphPad Prism, Dotmatics, Boston, MA, USA).
